# Incorporating and Addressing Testing Bias Within Estimates of Epidemic Dynamics for SARS-CoV-2

**DOI:** 10.1101/2020.05.02.20088120

**Authors:** Yasir Suhail, Junaid Afzal, Kshitiz

## Abstract

The disease burden of SARS-CoV-2 as measured by tests from various countries present varying estimates of infection and fatality rates. Models based on these acquired data may suffer from systematic errors and large estimation variances due to the biases associated with testing and lags between the infection and death counts. Here, we present an augmented compartment model to predict epidemic dynamics while explicitly modeling for the sampling bias involved in testing. Our simulations show that sampling biases in favor of patients with higher disease manifestation could significantly affect direct estimates of infection and fatality rates calculated from the numbers of confirmed cases and deaths, and serological testing can partially mitigate these biased estimates. We further recommend a strategy to obtain unbiased estimates, calculating the dependence of expected confidence on a randomized sample size, showing that relatively small sample sizes can provide statistically significant estimates for SARS-CoV-2 related death rates.

## INTRODUCTION

The spread of SARS-CoV-2 across the world has led to a significant disease burden with widespread health impact. While the search for a vaccine or a successful pharmaceutical agent continues, non-pharmaceutical interventions have been the only currently available recourse. Planning and implementing such interventions is intimately connected with epidemiological disease modeling and requires the estimation of key metrics such as the speed of infection spread, recovery and fatality rates, and kinetics related to the persistence or loss of acquired immunity. Early reports from the World Health Organization (WHO) stated a case fatality rate of over 3.8%^1^ for SARS-COV-2 as it was first detected in Wuhan, China and spread across the world. Subsequently, epidemiological modeling and projection, with its inherent estimation of infection, recovery, and fatality rates has become central to various institutional actors dealing with the management of epidemic. These studies and reports are, by necessity, ultimately based on reported numbers from tested patients, which were sampled by public health agencies in the countries where virus had started spreading^2–4^. However, there are wide differences across countries in the number of people who were tested, the availability of test kits, as well as the stratification of the population that were tested. With this criteria, the presentation of infection, recovery, and case fatality rate (CFR) based on current SARS-CoV-2 data is difficult to interpret^2–4^, masking the true extent and dynamics of the disease spread and ensuing fatality.

We were among the first to raise concerns regarding the accuracy of determined fatality rates based on two potentially important issues^5,6^. These issues deserve the attention of the scientific community involved in understanding the spread of the pandemic. One of these is the underlying spread of immunity. Reverse transcription polymerase chain reaction (RT-PCR), the commonly employed method to confirm the presence of SARS-COV-2, only informs about the live status of the virus in the population, and therefore may mask the percentage of people who contracted the virus and subsequently resolved the infection by acquired immunity^7^, if the disease has spread more than our current estimates suggest. SARS-COV-2 is known to induce a detectable antibody response following few days of infection^7,8^. An initial report suggested that a larger cohort of tested populations which were negative for an active viral load can now be regarded as having previously contracted the virus^9^, but the effect size found was smaller than could be statistically determined from error rate of the underlying test, besides concerns about the sampling bias during recruitment of the subjects.. Serological testing using a better study design by other researchers or public health agencies will not only shed more light on the true prevalence of COVID-19 resistance, but subsequent follow up of any COVID-19 immune individuals found therein will also answer very critical questions on the nature and persistence of COVID-19 immunity in the general population. Without sufficient attention to these questions, it will not be possible to model the chances of subsequent waves of COVID-19 spread in the future with much confidence. This in turn has grave implications for public health capacity planning and intervention decisions beyond the next few months.

Secondly, we highlight the bias within the sampling (testing for SARS-CoV-2 presence), which could potentially alter the estimates of both the infection and fatality rates. There have been multiple mathematical studies modeling the kinetics of disease spreading with and without social distancing interventions^10^. However, these are dependent on model parameters estimated from limited, and likely biased and non-uniform sampling, as indicated by large differences across countries in the rate of fatalities. The data collected across countries were collated for the objective of public health operations, identifying infected individuals and tracing their contacts etc., and for preparation of adequate health facilities. However, these approaches may introduce bias in testing for individuals presenting with symptoms, rendering models built on these data vulnerable to systematic sampling bias. This raises significant concerns regarding the accuracy of the estimates of fatality and morbidity rates, with far reaching consequences on capacity planning and policy making.

In this work, we present a new model to predict the dynamics of disease spread by augmenting the commonly employed SIRD compartment model by stratifying the infected population and introducing bias in their sampling. We show that biases within testing could have a significant effect on the estimation of the infection and fatality rates. We also accounted for testing by serological means and found that estimation of acquired immunity could partially mitigate the effect of testing biases. We also demonstrate that because death counts lag the infection rate, case fatality rates may underestimate the infection fatality rates.

However, large variance within the estimation of infection, as well as fatality suggested a strong need to determine the effect of bias within sampling itself. As the countries have introduced substantial social distancing, it has become difficult to predict if the disease has spread widely, or only in limited population niches. In which case the high estimation of fatality rates from positively tested patients in certain countries could either mean a high true fatality rate, or a high sampling bias wherein the more symptomatic patients with an enhanced likelihood of mortality are tested. Both these scenarios are theoretically plausible, and it is essential to estimate the true fatality rates associated with the epidemic to mount an appropriate response, or for an informed preparation^11^. Indeed, these numbers vary widely across different countries, resulting in large variations in suggested mortality rates^11^. In practice, kinetic models used to project the epidemic spread attempt to mitigate or sidestep the effects of sampling bias by various methods. Verity et al. give estimates for the infection fatality rates based on testing of foreign nationals repatriated from China^12^. While this sample may not have been directly biased with symptom severity, it is still likely to be highly correlated to age, health, and placement within social and physical contact networks, and therefore indirectly correlated with infection status and susceptibility to fatality. Other methods may fit on reported deaths, which are less likely to suffer from sampling bias, but could miss untested fatalities. We therefore propose that an unbiased randomly sampled testing study in a region with high fatality presents the best course to estimate the true fatality rates. Our calculations indicate that a reasonable unbiased testing sample can provide high confidence data to test the hypotheses of different fatality rates.

Together with our augmented compartmental model, our proposed scheme presents a coherent, statistically rigorous estimation method to determine infection and fatality rates, which is both cognizant of the testing bias in favor of the more symptomatic or severe patients, and given sufficient follow-up time, resistant to the underestimation bias due to lagging death counts.

## RESULTS

### An Augmented Compartment Model to Estimate Epidemic Dynamics Incorporating Testing Bias

We considered an augmentation of the currently prevalent models of epidemic dynamics to explicitly model the potential sampling bias within the tested populations for the specific disease, thus enabling the modeling of reported case numbers and the effects of different testing strategies. In the context of SARS-CoV-2 infection, this sampling bias is potentially large because data from different countries present very different fatality rates (**Figure 1A**). Indeed, directly measured case fatality rates (CFR) was proportional to how biased the sampling of positive tests were in each country (number of positive cases found per unit test conducted) (**Figure 1A**). We therefore attempted to incorporate biases involved in the stratification of populations tested for the active viral load, with an objective to study the effect of sample bias in the reported case numbers and any subsequent direct estimates of infection and fatality rate dynamics.

The most common model used to study epidemic dynamics is the SIRD compartment model, from which many derivatives have been designed. We decided to choose the simplest SIRD model to test whether testing bias could have an effect in the estimate of patients belonging to a given compartment. The basic SIRD compartmental model stratifies the population in 4 compartments: Susceptible (S), Infected (I), Recovered (R), and Dead (D). Movements of subpopulations from one compartment to the other are described by ordinary differential equations (ODEs). The parameters for each of these ODEs are the rate constants, β, describing the rate of infection, and γ, describing the rate of death (**Figure 1B**).

**Figure 1.**
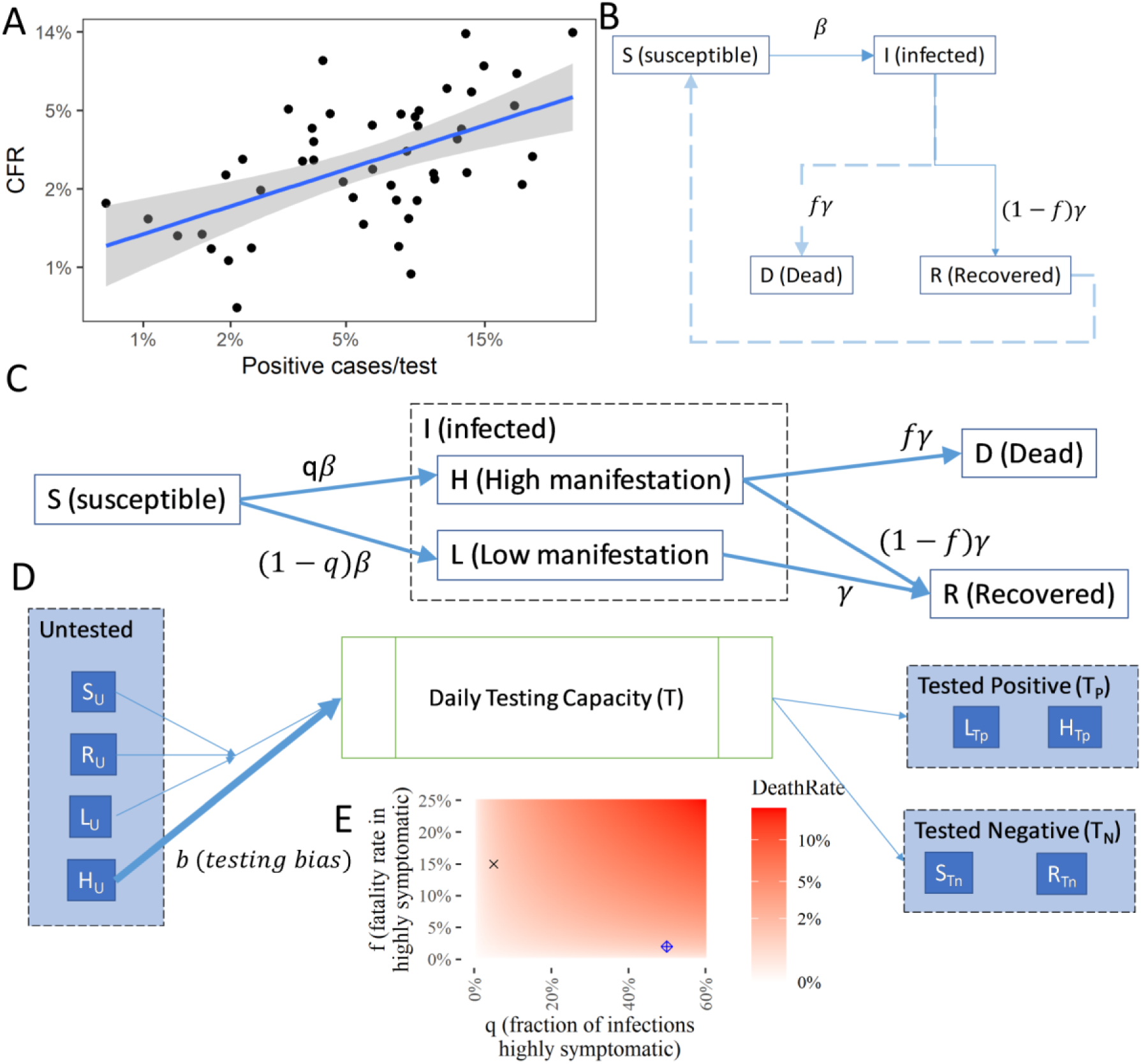
An Augmented Compartment Model to Predict Epidemic Dynamics with Testing Bias. **(A)** Regression of Case fatality rate, CFR (calculated as percentage of death in positively identified cases per country) against the percentage of positive cases identified among all tested per country show a linear regression; Each dot corresponds to a different country; Data obtained from our world in data for April 18, 2020; Blue line shows the fitted regression curve; Shaded area show the 95% confidence interval; R^2^ = 0.3567, p-value = 5.9e-6. **(B)** The basic SIRD compartmental model commonly used to model epidemic dynamics. Ordinary differential equations describe the movement of the population through the different compartments representing the susceptible, infected, recovered, and dead stages. The parameters are the rate constants for each term representing the transitions in the differential equation. **(C)** The augmented SIRD model by stratification of the infected population into H and L referring to high, and low manifestation of disease symptoms respectively; The factor q is the fraction of infected within H; We assume that high manifestation of disease leads to death in a fraction f of the individuals. (D) A simplified representation of the model of the testing policy; T tests are available per unit time; Untested alive individuals (U) are randomly selected in proportion to their numbers, but patients in H are selected with an increased bias b. Further compartments arising due to testing and movement at different stages are omitted here for clarity; Detailed equations in the Methods section. (E) Fraction f and q determine the true death rate; Two values with similar death rates chosen for simulations are marked.

In various countries, the initial tests for viral presence have been biased based on the severity of the disease manifestation, or weighted towards symptomatic patients. However, in certain situations, these biases could be present in other directions too, wherein patients with more likelihood of death are under-sampled. We therefore decided to introduce testing bias by stratifying the infected population based on the severity of disease manifestation. Specifically, we further stratified the infected compartment (I) into two other sub-compartments, H (high) and L (low) referring to the high or low symptomatic manifestation of the disease respectively. Although the transition from the S (susceptible) compartment to the infected (I) is driven by rate constant β, the factor *q* describes the fraction of the infected subpopulation manifesting a high symptomatic manifestation of the disease. We assumed that the fraction within L (low manifestation) die in miniscule rates, and nearly all deaths occur from the H fraction. This added sub-compartmentalization is a simple addition to the model, but we believe that if well-defined stratification could be measurably identified within the infected (I) compartment, more subcompartments should be added. These fractions could include people with known co-morbidities with a higher chance of fatality, or those with measurably high severity of disease manifestation.

We then superimposed upon our augmented compartment model a testing policy (**Figure 1C**). We assumed that T tests are available per unit time (kept constant for simulations below, but which could itself be a time varying function based on the availability of testing capabilities over time). The untested, and alive individuals are assumed to be randomly selected for testing in proportion to their numbers, but those with high disease manifestation (H) are selected with an increased bias b. In addition, patients who were tested as being negative for viral load at a previous time point, but presenting severe symptoms at the present time would also be selected with an increased bias *b*. The biased testing policy was implemented by splitting the compartments in our augmented SIRD model for the untested and the tested fractions (**Figure 1D**). True death rate would depend upon the factors q (fraction with high disease manifestation), and f (fraction dying within the H compartment) (**Figure 1E**). Details for the ODEs describing the transition through these compartments are provided in the Methods section.

### Testing Bias Strongly Affects the Direct Estimation of Infection Rate

We simulated our augmented compartmental model with testing bias and calculated the infection rate dynamics based on an active viral test (based on measurement of viral sequences), as well as based on a serological test (measuring if antibodies against the virus have been created). In our augmented model, the estimated infection rate is calculated as a ratio of those tested positive, and all tested population within a given time frame. Here, the testing bias is reflected within the sampling of stratified populations, H and L in the infected (I) compartment.

Assuming no errors within the tests (sequence based, or serological), we found that testing bias had a profound effect on the naïve estimation of infection rate based on active viral test, the most commonly employed tests (**Figure 2A**). In contrast, estimation of the contraction rate was much less affected by the bias, largely because the immunologically recovered population as a fraction increases as time progresses (**Figure 2B**). Our simulation provides a strong argument in favor of serological testing beyond the obvious argument of their capability to correctly assign the compartment of recovered (R) fraction to the population which contracts the disease but tests negative. That the testing bias could be substantially mitigated in the estimation of contraction rate by serological test is a strong argument in favor of serological testing, although these tests are unlikely to be available in the initial spread of the epidemic. Nevertheless, our augmented model will allow estimation of the effect of biased sampling itself in predicting disease dynamics, and underlines the importance of unbiased sampling to predict estimates reflecting reality.

**Figure 2.**
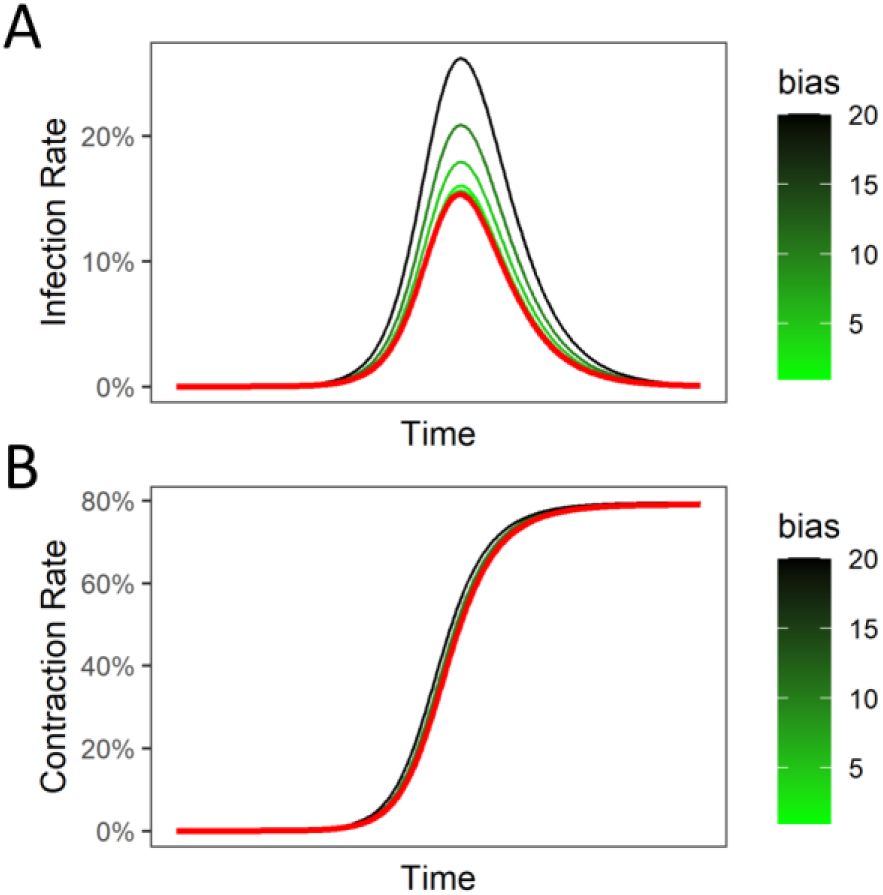
Influence of testing (sampling) bias for symptomatic patients is high for estimation of infection rate measured by active-viral tests, but mitigated in serological tests for acquired immunity. **(A)** The infection rates (fraction of population with an active infection) estimated from PCR (or sequencing) based strategy to measure active viral load; Red curve shows the true infection rate; Estimation of infection rates with different biases for H-compartment patients (those with higher disease manifestation) shown in green-black lines; Also shown are fractions calculated in equations. (B) Estimation of contraction rates (fraction of population that has contracted the virus at a previous time, are either infected or recovered) from a serological test; Red line is true contraction rate; Green-black lines are estimated contraction rates with different sampling biases for patients with higher disease manifestation.

### Testing Bias Influence True Fatality Rates and Case Fatality Rates in Time-Dependent Manner

Since the initial report of case fatality rate of 3.8% from Wuhan China by WHO, there has been a substantial variance in the listed death rate among nations. Case fatality rate is calculated as the ratio of number of deaths measured and number of positive cases^2^. Since death is a lag indicator, case fatality rate would asymptotically reach the more conservative death rate, which is the ratio of deaths and a sum of those who died or recovered. We, therefore, considered the latter death rate and tested the effect of sampling bias upon its estimation. We found that sampling bias can linearly affect the estimation of death rate (**Figure 3A**). If the fraction of population with high manifestation of disease is high (and therefore the inherent bias of sampling is low), then trivially the effect of bias is somewhat mitigated (**Figure 3B**). Our data strongly underlines the importance of incorporating bias in testing itself as a key parameter to model epidemic dynamics, because key predicted metrics, including death rate, are substantially affected by these biases.

**Figure 3.**
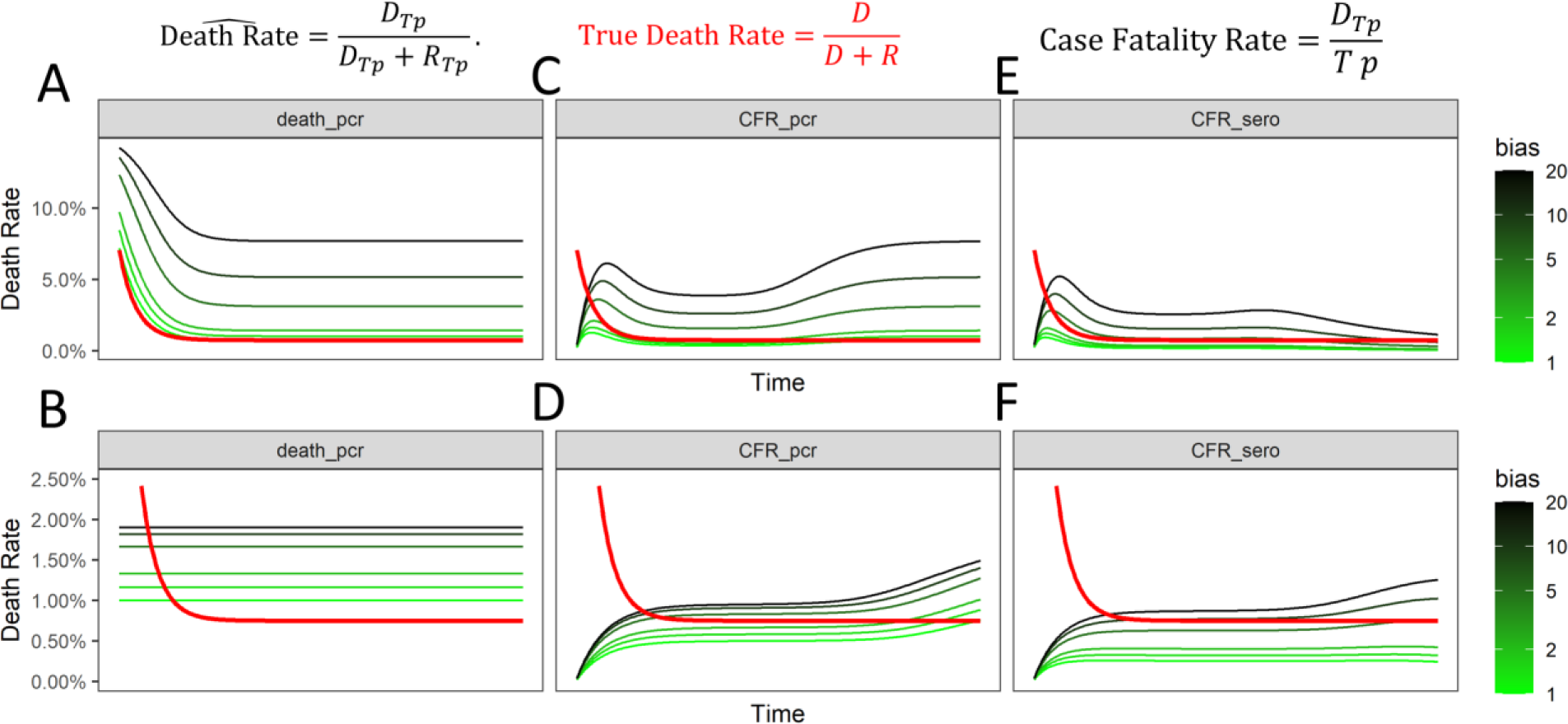
Estimates of Fatality Rates are influenced by biased sampling of patients in a disease manifestation dependent manner. **(A-B)** Estimates of death rate (percentage of fatalities within confirmed recovered and dead patients) from a biased sampling strategy for patients with high disease manifestation at different fractions resulting in similar true death rates: (A) 5% of the infected population with 15% death rate, and (B) 50% of the infected population with 2% death rate; Red lines refer to true death rate and green-black lines refer to biased estimates of death rates. (C-D) Estimates of case fatality rate (percentage of confirmed fatalities within positively tested population) from a biased sampling strategy as in A, and B for different fractions of population in H and L manifestation, as in A, and B. (E-F) Estimates of case fatality rate measured with serological testing from a biased sampling strategy as in A, and B for different fraction of populations in H and L manifestation, as in A, and B.

We then tested how testing bias would affect the case fatality rate (ratio of dead to the number of positive cases). We found that sampling bias indeed resulted in large effects in CFR estimates, but crucially, these effects reduce as the infection reaches its peak, and then amplify as the infections subside within the population (**Figure 3C**). When compared to the true death rate, CFR initially underestimates the death rate, and then overestimates the rate in a bias dependent manner. CFR, as is calculated here, has two opposing biases inherent in it. In the initial part of the pandemic, and for testing biases below a threshold, it underestimates the true death rate because the growth of the infections happens earlier than the growth in the number of death. In other words, while the pandemic is growing, the number of deaths always lag, and the number of infections at a particular time are some multiplicative factor larger than the corresponding infections that existed when the currently dead were infected. This leads to estimation of an overly optimistic CFR. On the other hand, for later and waning stages of the epidemic, and the testing biases being greater than a threshold, the CFR leads to an overly pessimistic number compared to the real death rate. This is due to a greater prevalence of the severely ill patients counted among the cases. Indeed, if the fraction of population with a high disease manifestation (H) are changed and correspondingly the death rate adjusted to keep the true death rate the same, then CFR can underestimate the true death rate for a longer duration of the pandemic (**Figure 3D**). The direction of the bias in the CFR depends on the pandemic kinetics *(β, γ)* and the testing bias.

However, if CFR were to be measured using serological tests, thereby counting the recovered population as being previously infected, the effect of bias on estimation of fatality rate is mitigated (**Figure 3E-F**). Crucially, our calculations argue that for all metrics of naïve estimation of fatality rates, contribution of testing bias is substantial, and attempts made to measure the bias itself, and be accounted for.

### Randomized Unbiased Serological Sampling of Widely Infected Population is Necessary to Determine True Fatality Rates

Our augmented model demonstrated that sampling bias could play a significant role in the direct estimates of both the infection and the fatality rates, and may partially explain the large variance across the death rates reported across countries, as well as in epidemic prediction models. We therefore propose that a random sampling of a population that has suffered a large infection load should be utilized to estimate the true infection, recovery, and fatality rates. In addition, as serological tests are becoming available to detect the antibodies against SARS-CoV-2 specific antigen, it is possible to identify individuals who have developed immunity against the virus but may not necessarily test positive owing to reduced viral load. We believe it is necessary to test if the wider population in an area with high death numbers is a consequence of (i) a wider spread of the disease and a smaller death rate, or (ii) a larger death rate in a smaller subpopulation that has contracted the virus. In order to minimize the variance of the infection death rate, this random testing is suggested to be conducted among a population wherein the infection is understood to have spread widely.

We propose using census, tax ID, or driving licenses in a given area as an unbiased identifier of a sampling set, upon which the serological test and PCR (or pooled next generation sequencing-NGS based tests) could be conducted. We calculated the sample sizes required to gain an accurate estimate of the community infection rates and the infection fatality rates. A random selection of individuals will provide estimates without systematic biases; therefore, the appropriate measure of accuracy need only be concerned with the variance of the estimates. In the following, we have chosen to frame this in terms of mostly confidence intervals and hypothesis testing.

An initial calculation expectedly suggested that with low infection rates, attaining a 5% error of estimation for the infection rate would require a moderate sample size. In contrast, if the real infection rate is higher (closer to 50%), expectedly, a smaller sample set will be sufficient for an accurate estimate of infection rate (**Figure 4**). Therefore, in the present scenario, an example of an ideal location where such tests could be performed with a limited number of sample size (approximately 10,000) is New York City, where the deaths have rapidly climbed up in the last week of March 2020.

**Figure 4.**
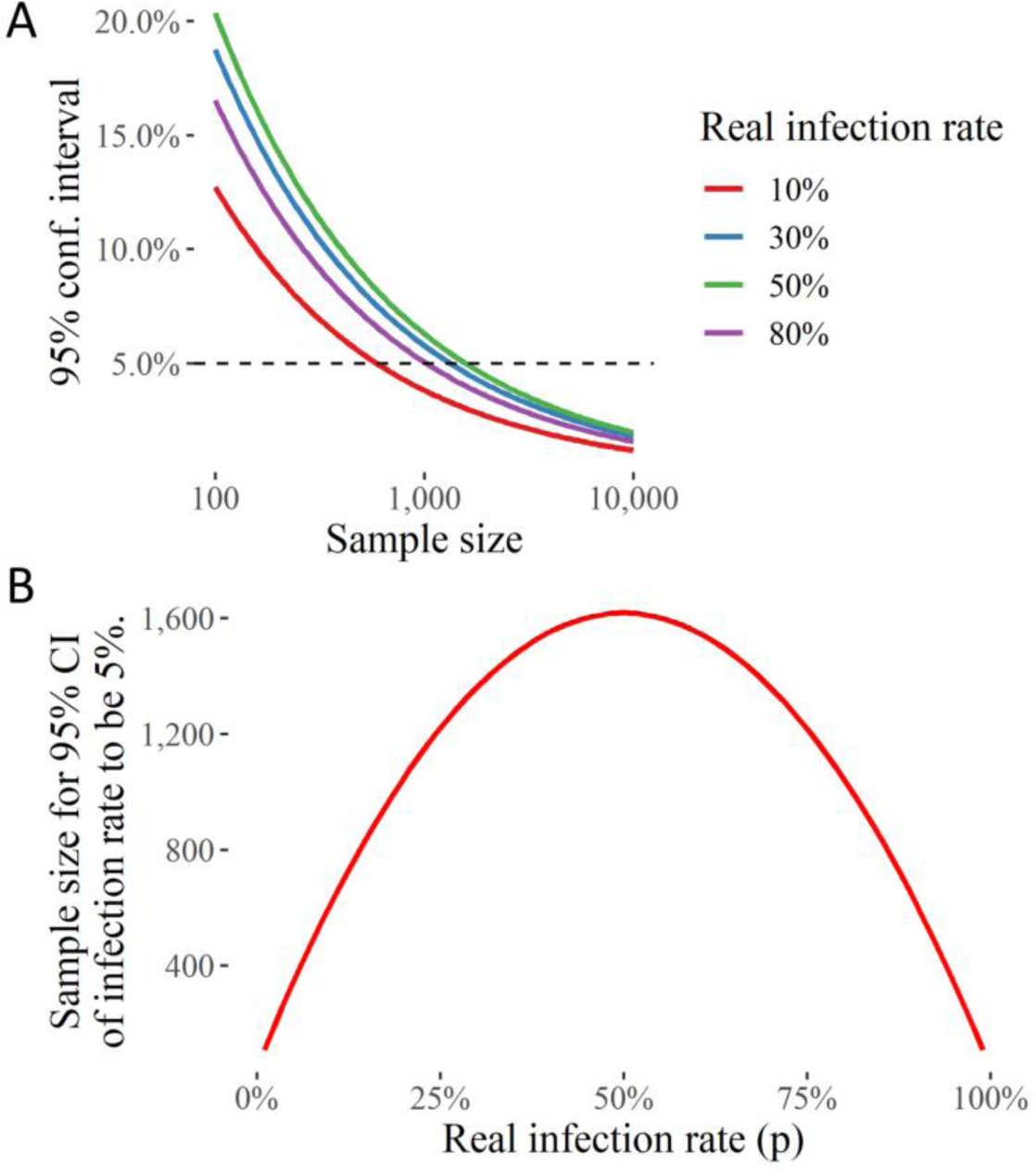
Randomized Testing Strategy for estimation of SARS-CoV-2 infection rate in an area with a high infection rate. (A) The uncertainty (in terms of the 95% confidence interval) in the estimate of the fraction of population with SARS-COV-2 (infection rate) with different sample sizes. (B) The sample size needed for infection rate 95% confidence interval to be 5%.

Estimating the mortality or infection fatality rate requires another probability to be multiplied to the estimate of infection rate within a sample population. A calculation of the sample size required for a 95% confidence interval indicates that even for a potentially highly infected population, like in NYC, it may require a very large sample size to accurately determine the true fatality rate (**Figure 5A**). This may be one reason why countries have resorted to large sampling to obtain data for true fatality rate. However, biased and non-random sampling renders these data difficult to interpret to estimate the fatality rates.

We therefore propose to instead test the hypotheses that the true fatality rate is higher than a given value, which would be rejected if the upper limit of the 95% confidence interval is lower than the said value. Calculating these sample sizes with the statistically significant 95% confidence interval, we found that a relatively much smaller sample size would be sufficient to estimate if the true fatality rate is below or higher than a given percentage (**Figure 5B**). Our calculations indicate that for a sample with 50% infection rate, a sample size of 1,000 may be sufficient to identify if fatalities are much lower than 1%, while for a sample with 25% infection rate, it may be below 10,000 — a logistically achievable size to determine a crucial parameter.

**Figure 5.**
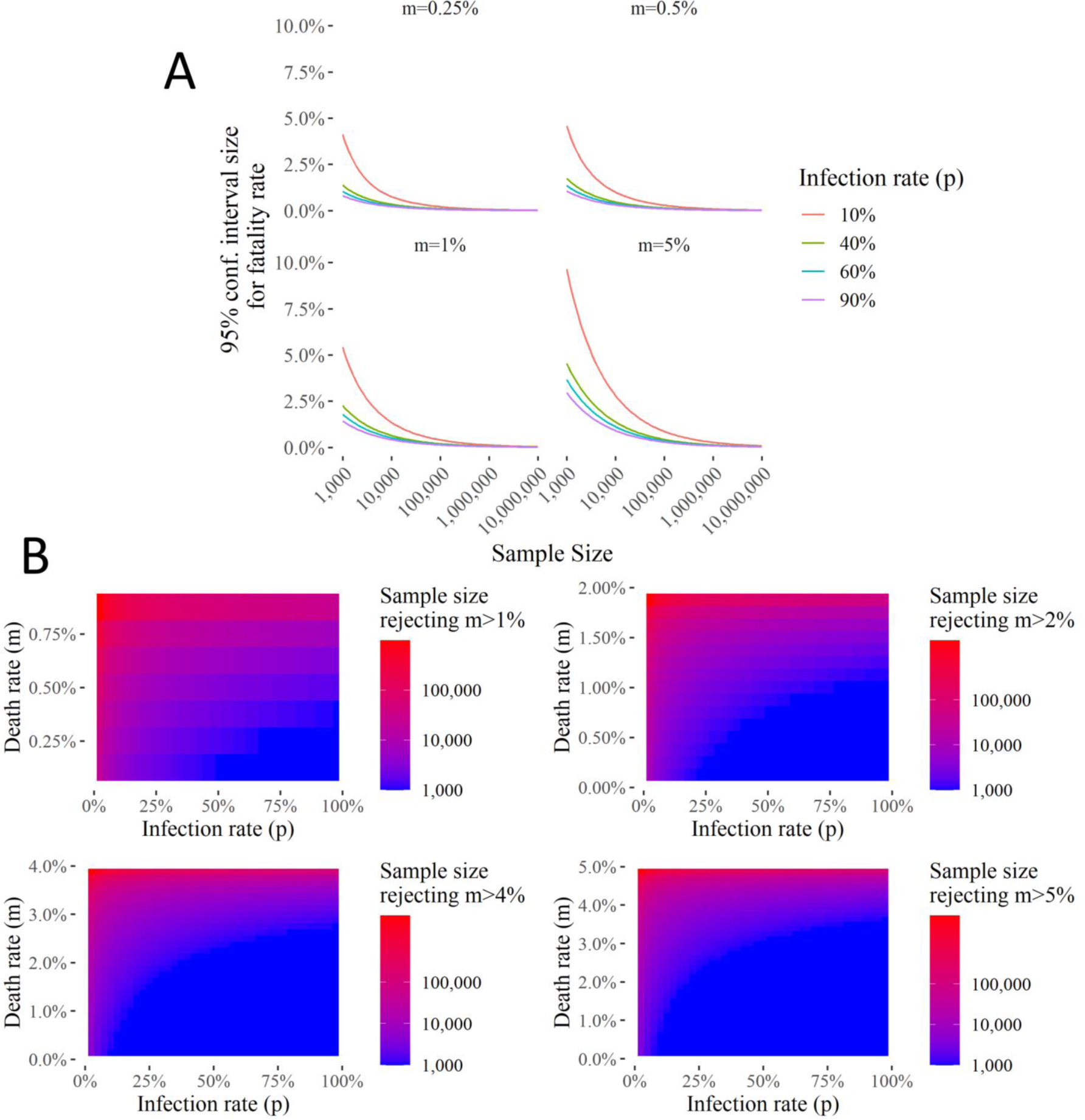
Randomized Testing Strategy for estimation of fatalities, and for estimate of death rate below a given percentage in areas with different infection and fatality rate. **(A)** 95% Confidence Interval size for the death rate given a sample size (S, x-axis), infection rate (p, line colors), and the real death rate (m, subplot panel). **(B)** Sample sizes needed to reject hypotheses that death rate > than 1, 2, 4, or 5% of the infected population.

### Continuous Sampling of a Selected Cohort can Provide Useful Dynamics on Acquirement of Immunity

The availability of a serological test, if applied using a random and unbiased sampling strategy could allow the identification of a key subset of people who have developed immunity, but do not carry the infectious disease burden. However, it is usually not possible to have antibody tests available at the onset of a disease, and a rapidly spreading pandemic may make it difficult to gear policies based on an accurate assessment of the development of herd immunity. In contrast, the recent development of genomic amplification or sequencing technologies has made it possible to prepare rapidly deployable tests to assess active infectious loads. We therefore propose to use a continuous sampling of a representative unbiased cohort on a weekly basis to determine the initial onset of infection, the rate of its spread, development of immunity, and eventually the ensuing aftermath of the infection. Indeed, as we showed, for very small infection rate, a larger sample may be required. However, this concern is easily addressable by pooled sequencing (NGS), which can be used to determine rare onset, mutagenesis, and characterization of infections^13–15^, and if sufficient signal for infection is found, then the continuous sampling be used for that cohort. The dynamics of readout (of active viral load) in a fixed sample set will allow an accurate estimation of the development of immunity and its dynamics in a given population.

## DISCUSSION

The wide, and constantly updated, estimates of key metrics of the disease, including fatality and recovery rates associated with SARS-COV-2 raise important questions about the quality of our public health scientific inquiry. Additionally, the effects of the public health and economic policies adopted around the world on the socio-economically and politically vulnerable sections of the population has so far received insufficient attention. This is the most severe global health crisis to have inflicted humanity within this generation, although its true impact has still not been understood completely. Indeed, even after months of its spread, there is a large variation in the estimate of infection, recovery, and fatality rates. Crucially, an accurate estimate of the true dynamics and infection, recovery, and fatality rates is necessary for the scientific inquiry into the disease from a systems perspective, operations planning and to advocate for an apt public health policy. We show that direct estimates of these parameters lack in this respect methodologically. Testing of the general population has been justifiably biased towards symptomatic patients, since it is driven by the desire to identify, care for, or safely isolate vulnerable populations rather than to estimate accurate metrics. This has resulted in the biased sampling that is not perfectly suited for modeling of the epidemic and calculation of key metrics.

In this work, we attempted to systematically model this crucial issue by incorporating the testing bias within the compartmental model of epidemic dynamics. Our model also includes tests for active viral load, as well as for those who have developed immunity, along with the sampling bias in testing. We believe that our proposed augmentation not only provides a systematic basis to ascertain the effect of testing policies in estimates of epidemic dynamics, but also demonstrates that biased sampling may substantially influence epidemic projections if metrics are naïvely calculated only reported case numbers.

Another problem that we demonstrate with the directly calculated case fatality rates is that the counts of deaths lag the infection rates, and therefore while the pandemic is growing, the case fatality rates underestimate the infection fatality rates. While the bias introduced due to this lag is in the opposite direction to that introduced by the sampling bias, the result only makes the situation worse. In terms of statistical theory, the case fatality rate is a large variance, biased estimate with an unknown direction of bias. Case fatality rate estimates calculated for countries with proportionately very extensive testing such as Iceland have been optimistically cited as the true infection fatality rates. However, as some sources of systematic errors are mitigated in extensive testing, the effect of the lagging death counts will proportionately become more important. Therefore, until we arrive towards the end of the pandemic, these optimistic case fatality rates may be more optimistic than the reality.

Although much data is collected on the number of cases, the ensuing deaths, and those that have recovered, the naïve interpretation of fatality and infection rates from non-uniform data across countries may be fraught with substantial inherent problems. Therefore, we recommend a limited, unbiased, random uniform sampling of population to test hypotheses of fatality rates. We also propose a method to continually monitor a static sample set to estimate the onset, and dynamics of disease spread, acquired immunity, and ensuing morbidity and fatalities associated with an infectious spread. As a recent example, a large number of deaths in New York City could be explained either by (i) a high fatality rate in a small population contracting the virus, or (ii) a rapid spread of the virus which has resulted in large number of people to develop immunity with a smaller percentage succumbing to the viral infection. In order to distinguish between the two widely varying scenarios, the most direct method with the least amount of statistical assumptions, would be to serologically test a limited, random sample of individuals. This should occur in addition to any surveillance methods currently being employed that are independently needed for targeted medical care and public health interventions.

## METHODS

### SIRD model with disease stratification into high and low symptomatic populations

First, we augment the canonical SIRD model by stratifying the infected population into high (H) and low (L) symptom populations. The differential equations for disease progression can be written as

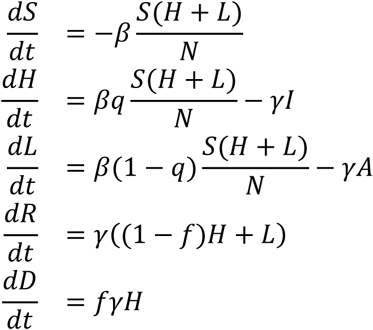

where *S* stands for the susceptible population, *R* for the recovered population, *D* for dead, and *H + L* for the total infections. *β* is, according to convention the infection rate constant, and *γ* the recovery rate constant, 0 ≤ *q ≤* 1, is the fraction of infections that develop the *H* manifestation of the disease, and 0 ≤ *f* ≤ *1* is the fraction of the *H* disease population that dies from it.

### Augmented SIRD model with testing

We go one step further that model the testing for the infection conducted as part of a surveillance program, under the following assumptions:

1. Both uninfected and infected individuals can be tested in a surveillance program, with different probabilities.
2. The amount of surveillance testing capacity is limited to *T* tests per unit time (day).
3. Uninfected people who are tested, and test negative aren’t tested again, unless they show significant symptoms *(H)* at some point later.
4. Once an individual has tested positive, they are a confirmed case, and any further testing etc. as part of the care program is not counted in this model since such testing will not change the confirmed case numbers, and isn’t assumed to come from the surveillance testing capacity.

In total, we have the following states

1. *S_U_*, the untested susceptible population,
2. *H_U_*, the untested infected highly symptomatic,
3. *L_U_*, the untested infected with none or low levels of symptoms,
4. *R_U_*, the untested recovered population,
5. *D_U_*, the population that died from the disease without being tested
6. *S_Tn_*, the susceptible population that has been tested, and obviously tested negative,
7. *H_Tn_*, the highly symptomatic infected population that was earlier tested negative during the susceptible phase (but might be tested in the future during infection)
8. *L_Tn_*, the low symptom population that was only tested while susceptible, and therefore tested negative at that time,
9. *H_Tp_*, the high symptom infected population that was tested while in the infected stage, and hence tested positive,
10. *L_Tp_*, the low symptom infected population that was tested while in the infected stage, and hence tested positive,
11. *R_Tn_*, the recovered population that was tested in only the susceptible or recovered stages, and hence tested negative,
12. *R_Tp_*, the recovered population that was tested in the infected population, and hence tested positive,
13. *D_Tn_*, the deaths due to the epidemic, that were tested negative, and
14. *D_Tp_*, the deaths due to the epidemic, that were tested positive.

The dynamics from the state transitions is written as

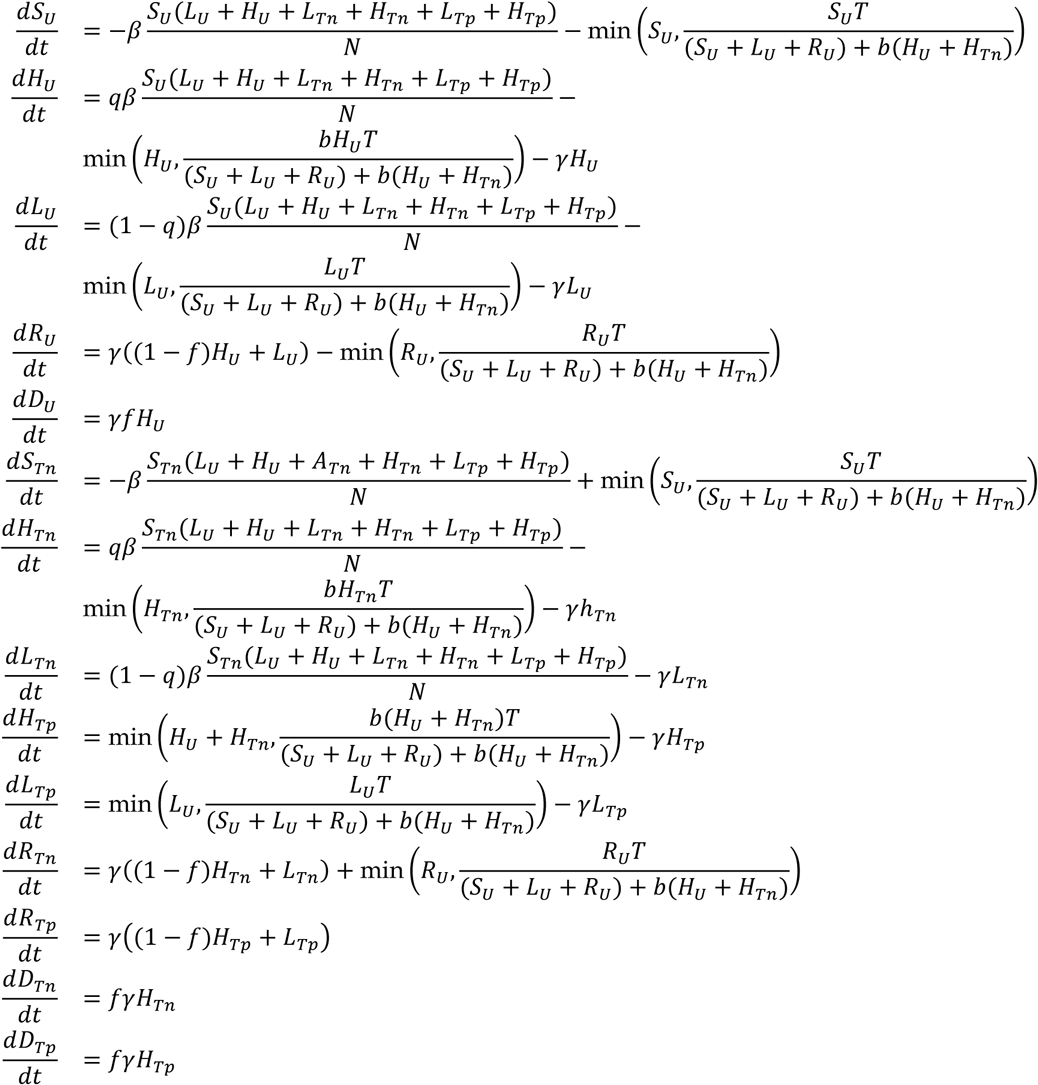

In addition, the cumulative number of positive and negative tests can be calculated as

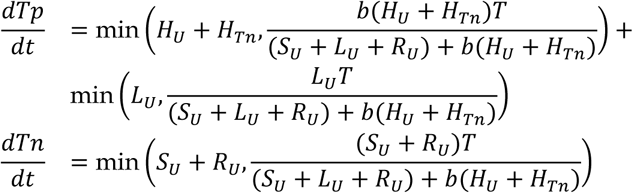

In case serological tests are done, the cumulative number of positive and negative tests can be calculated as

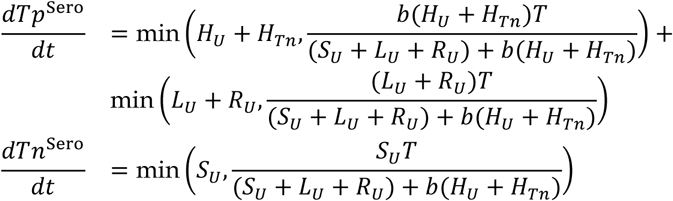

### Estimates of infection and death rates

Using the testing results, the conventional estimate of the infection rate as currently being reported would simply be the fraction of positive test cases found in a time period

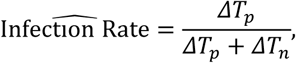

and the cumulative death rate estimate would be the calculated from the number of people who died from the pandemic versus those recovered

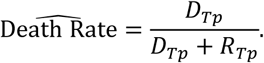

The case fatality rate, as it is being currently being defined is

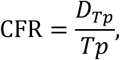

which would change to

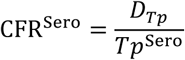

if we use serological testing.

Instead, the true infection rate in the population would simply be the fraction of the population with any kind of infection

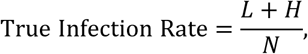

and the true death rate would be simply the total number of people who died of the disease versus the total that died or recovered

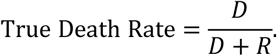

### Calculating the variance of estimates for unbiased random sampling

Suppose the fraction of population that has contracted SARS-CoV-2 detectable by a serological test (the infection rate) is *p*. In addition, assume that within those with SARS-CoV-2, a fraction *m* have died or die within the study time-frame. Therefore, in sampling a random sample of *S* samples, we expect to find *Sp* positive cases, and *Spm* deaths. In terms of the sampled numbers, if we find *C* positive cases out of a total *S* sample size and *D* deaths, the estimates of the infection rate will be *p = C/S* and the estimate of mortality rate *m = D/C*. These are unbiased estimates, and their conservative, guaranteed confidence interval can be calculated from the Clopper-Pearson interval^16^.

## Data Availability

No new patient data is generated, but all models will be made available upon publication in peer reviewed journals.

## DECLARATION

No data is collected for this manuscript, and therefore no consent was required. There are no competing interests declared. Funding was provided by the UConn Health Startup Funds. The authors listed have contributed towards the manuscript in the following aspects: K and YS conceived the idea, YS created the statistical groundwork and generated the figures, JA provided the medical rationale and helped in writing the manuscript with all other authors.

### Biography of First Author

Yasir Suhail is a Bioinformatics Statistician, having completed his B.Tech. in Electrical Engineering at IIT Delhi, and Ph.D. in Biomedical Engineering at the Johns Hopkins School of Medicine with a concentration in Bioinformatics and Statistics.

